# Can women’s reports of client exit interviews be used to measure and track progress of antenatal care quality? Evidence from a facility assessment census in Malawi

**DOI:** 10.1101/2022.09.03.22279523

**Authors:** Martina Mchenga, Ronelle Burger, Dieter von Fintel

**Affiliations:** Graduate School Of Economic Sciences and Management, Stellenbosch University, Western Cape, South Africa; Economics Department, Stellenbosch University, South Africa

## Abstract

**Introduction:** Exit interviews given their shorter recall period unlike household surveys provides an opportunity to capture routine performance and level of service quality at the facility level. However, very few studies exist validating whether women’s reports in exit interviews can reliably be used to measure quality of care. This study contributes to the sparse literature by examining the validity and reliability of women’s report of selected antenatal care (ANC) interventions in the exit interviews in Malawi.

**Methods:** Using the 2013-2014 Malawi service provision facility census, we compared women’s reports at exit interviews of ANC received with reports through direct observation by a trained third-party. Six indicators of ANC common in both direct observation and client exit tool were identified. These include; whether provider prescribed or gave fansidar for malaria prevention; whether provider prescribed or gave iron and folic tablets; whether provider explained side effects of iron and folic tablets; whether provider discussed importance of good nutrition during pregnancy; whether provider discussed delivery preparation and whether provider discussed pregnancy related complications. We assessed indicator accuracy by calculating sensitivity, specificity, area under the receiver operating characteristic curve (AUC) and inflation factor (IF). Indicators considered to have both high individual accuracy (an AUC value of 0.70 or greater) and low population-level bias (0.75<IF<1.25) were considered to have acceptable validity. To measure agreement, we calculating Kappa coefficient (κ) and the prevalence adjusted bias adjusted kappa (PABAK). Both κ and PABAK values were interpreted as follows: almost perfect (>0.80), substantial (0.61–0.80), moderate (0.41–0.60), fair (0.21–0.40), slight (0.00–0.20) and poor (<0.00). Using chi-squared tests we also examined factors that are associated with high agreement between exit interview reports and direct observations.

**Results:** for both validity and agreement measures, our findings show that women’s self-reports in the exit interviews presented overall higher accuracy and agreement for indicators related to concrete, observable interventions. For example, indicators which met accuracy criteria reflected those to do with medical prescription. In contrast, indicators related to counselling or advice given, performed less reliably. The results also show that age, primiparous status, number of antenatal visit and type of health provider were associated with high level of agreement.

**Conclusion:** In the context of calls for enhanced measurement of the components that lead to effective coverage, study findings such as these suggest that careful consideration of the type of information women are asked to recall is needed and also the timing is important. While household survey programmes such as the DHS and MICS are frequently relied on as data sources for measuring intervention coverage, triangulation of such findings with other data sources such as client exit interviews is important.

## Introduction

In low and middle-income countries (LMICs), relatively high antenatal care (ANC) coverage still continues to co-exist with high maternal and neonatal mortality rates (1, 2,3). In Malawi, for example, 95% of women attend ANC at least once, while maternal mortality is estimated at 497 per 100,000 live births and neonatal mortality at 27 deaths per 1,000 live births (4). This weak relationship between ANC use and maternal and new born survival has motivated a shift in the focus from quantity of care to content and quality of care provided (3, 5). This shift to focusing on quality of care therefore, underscores the need to monitor and track progress in the coverage and quality of care given (6). Hence, accurate and quality data on coverage and quality indicators is critical for global monitoring of trends and at national levels to provide actionable information to achieve desired health outcomes (7).

In Malawi, similar to most Low and Middle Income Countries (LMICs) household surveys such as the Demographic and Health Surveys (DHS) and the Multiple Indicator Cluster Surveys (MICS) are mostly used to track both coverage and quality of maternal and newborn services including antenatal care. However, the major limitation with household surveys is the longer recall period (up to 5 years) (4), therefore, their reliability also depends on whether the woman is able to provide a consistent recollection of what happened in the past, and that a woman’s current characteristics or conditions do not significantly influence her ability to accurately remember what happened (anchoring) (8). For example, if a child had satisfactory birthweight, and is growing up well, mothers may be more likely to recall quality of care in a positive light, leading to upward bias in quality measurement. Moreover, evidence shows that women’s recall of maternal and immediate postnatal interventions changes over time and that the longer the period, the less accurate the recall (7, 9), thereby questioning the validity of using household surveys with longer recall for monitoring and tracking quality of MNCH interventions.

Given the short falls of household surveys, facility level data provides a reliable alternative. There are a number of methods that are used to collect service coverage and quality of care information at the facility level including review of medical records (10); direct observation of clinical consultations by an expert; client exit interviews (11); and mystery patients (12). Of these, medical records have been shown to be the most reliable method of collecting data on service coverage and quality of health care (10, 13) as they allow retrospective assessment of routine provider performance. However, in LMICs including in Malawi, the use of medical records is often of little use due to incomplete, inconsistent or even non-existent records, particularly at public facilities (14). As such, service coverage or quality of care estimates at facility level are either collected using direct observation or client exit interviews.

Direct observation involves recordings of the provider’s actions during a consultation by an independent observer, is mostly considered a gold standard (15). Franco et al. (16) argued that information derived from direct observation, when the independent observer simultaneously records the providers actions using structured checklist to assess whether he/she is following a set of guidelines, has the potential to provide one of the most complete and reliable pictures of what providers do. Unlike household surveys, client exit interviews are conducted immediately after a consultation and therefore, provides an opportunity to capture routine performance and level of service quality (17). Currently, limited research exist that assess the client’s ability in exit interviews to recall with accuracy the services they received and the quality during a consultation. Given that exit interviews just like household interviews are less costly but provide a better alternative of collecting data given their shorter recall period, such a study is important.

Bessinger and Bertrand (18) conducted a study in Ecuador, Uganda and Zimbabwe to examined the comparability of reports from direct observations and exit interviews at health facilities. The authors found that for the majority of indicators, agreement was good to excellent and was relatively higher on the indicators of interpersonal relationships but lower on those related to counselling. A similar finding was also reported by McCarthy et al (19) who found that women’s reports in exit interviews of antenatal and postnatal care received in Bangladesh, Cambodia and Kenya had higher validity for indicators related to concrete, observable actions, as opposed to information or advice given. Assaf et al (20) examined the agreement of direct observations and client interviews in Malawi, Haiti and Senegal and also found overall low agreement in counselling topics related to antenatal care. As far as we know, this is the only study that includes Malawi, however, the study only focuses on counselling components of maternal and child health services. Our study, provides an extension to this study by focusing on ANC interventions in Malawi and includes other aspects besides counselling.

Given the renewed focus on measurement of quality-adjusted coverage, as well as the limited number of studies that have sought to assess self-reported ANC interventions, additional validation work is warranted. The present study aims to extend research findings to date by assessing the validity of a set of antenatal indicators that reflect a range of recommended interventions and counselling procedures in Malawi. The study also uncover factors that are associated with agreement between women’s self-reports and expert direct observation reports.

## Data and methods

### Study design

The study used secondary data from the 2013-2014 Malawi Service Provision Assessment (MSPA) collected by the Ministry of Health with the financial support from the United States Agency for International Development (USAID) (21). The 2013-14 MSPA was designed to be a census of all formal-sector health facilities in Malawi with a master list of 1,060 facilities provided by the Central Monitoring and Evaluation Division (CMED) of the Malawi Ministry of Health. Of the 1,060 formal health facilities that were visited during the assessment, 83 facilities were permanently closed, unreachable, duplicates of other facilities, or refused to participate (21). Data is therefore available for a total of 977 facilities.

The survey used four questionnaires to collect data on various aspects of quality of care. These include: (1) an inventory questionnaire, examined the availability of services and features of the facility; (2) the health worker interview, collected information from 8-15 selected health workers on their duties, training, and demographic characteristics; (3) observation protocol by a health worker with expert knowledge and experience, assessed providers’ adherence to clinical guidelines during consultations. The number of observations ranged between 5-15 consultations per provider and per service depending on the size of the facility. Facilities where direct observations were conducted were chosen at random to avoid sample selection bias. And (4) exit interviews, in which a client whose visit was observed provided their perceptions of the visit, recalled the services received and provided demographic information.

For this analysis our focus was on comparing reports from observation protocol by experts and women’s exit interviews. In this analysis, data from direct observations served as a gold standard and reference. Since direct observation and client exit interviews were conducted only in selected facilities that offer ANC, we conducted descriptive analyses to compare characteristics of facilities with observation data and those without (see Appendix 1) (22). We found that facilities without observation data were mostly lower-level facilities offering primary healthcare (non-hospitals) with ANC services offered for less than five days per week.

### Sample size

The total number of facilities where ANC direct observations were conducted was 412. A total of 2105 women were available on the day of assessment at the selected facilities, however, 2068 were both observed and interviewed after the consultation representing 98% response rate. In ANC consultations, experts observed whether health workers conducted routine tests and prevention procedures outlined in ANC guidelines. And in the exit interviews, women were asked to recall about the services they received, their perception about the services as well as other socio-demographic characteristics such as education level, number of ANC visit, whether it was the first pregnancy among others.

### Variables of interest

Indicators included in this study were selected based on two criteria: (1) interventions that are outlined in both the WHO ANC guidelines (23) and Malawi ANC guidelines (24) and (2) availability of the indicators in both the observation guide and the exit interview questionnaires. We identified the following six indicators in both questionnaires; whether provider prescribed or gave fansidar for malaria prevention; whether provider prescribed or gave iron and folic tablets; whether provider explained side effects of iron and folic tablets; whether provider discussed importance of good nutrition during pregnancy; whether provider discussed delivery preparation and whether provider discussed pregnancy related complications.

Binary variables were constructed for each element to determine whether women received any of the mentioned ANC elements using each of the two data sources. For direct observations, we coded the indicator for provision of an ANC service as 1 if the observer noted that the provider provided the service and 0 otherwise. Cases with missing information and where the provider was not observed providing the service were recoded as 0 and assumed not to have occurred (22). On the other hand, in the client exit interviews, women were asked whether they had received a particular ANC service during the current visit, both the current and the past visit or during the last visit only. For comparability with direct observations, we only coded client exit variables as 1 if a woman reported to have either received any of the ANC elements in the current visit, or in both the current and the previous visit. The validity measures were conducted at a client level.

For validity studies, adequate sample size is important to ensure precise estimates of specificity and sensitivity. In our study, given that the assessed indicators were health promoting and prevention focused and should be nearly universal and following McCarthy et al (19), we anticipated the prevalence of indicators to range from 50% to 80%. Since the interviews were conducted on the same day of the consultation and recall was immediate, we assumed levels of moderate to high sensitivity (60%–70%) and specificity (70%–80%). Using Buderer’s formula (25), the sample size for anticipated sensitivity and specificity levels was calculated a sample size of 400 women to be sufficient. Therefore, a sample of 2068 is more than enough to give us precise validity estimates.

## Analytical Methods

A two by two table was constructed to calculate to calculate sensitivity (the true positive rate) and specificity (true negative rate) for each indicator for validity measures. We estimated the area under the receiver operating characteristic curve (AUC) and its corresponding 95% CI following a binomial distribution. The AUC can be interpreted as ‘the average sensitivity across all possible specificities (26). An AUC of 0.5 indicates an uninformative test and an AUC of 1.0 represents perfect accuracy (100% sensitivity and 100% specificity) (26). Following McCarthy et al (19), an AUC value of 0.7 or higher was used as the cut-off criteria for high individual-level reporting accuracy. To assess population-level validity for each indicator, we calculated the degree to which an indicator would be overestimated or underestimated in the exit interviews using the inflation factor (IF). Specifically, the IF is the ratio of the indicator’s estimated population-based survey prevalence to the indicator’s ‘true’ (observed) prevalence. The population-based survey prevalence (Pr) is estimated as follows;

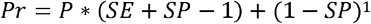

Where SE is sensitivity and SP is the specificity and P is the indicator’s true observed prevalence. We used an IF cut-off between 0.75 and 1.25 as the benchmark for low population-level bias (27).

On the other hand, reliability measures or agreement between direct observation and women’s reports in the exit interviews were calculated using Kappa coefficient (κ) with corresponding 95% confidence interviews (95% CI). Kappa coefficient (κ) is the proportion of agreement beyond expected chance agreement. Although Kappa remains the most widely used measure of agreement, several authors have pointed out data patterns that produce a κ with paradoxical results. For example, if the agreements bunch up in one of the agreement cells (prevalence) or disagreements bunch up in one of disagreement cells (bias), then the κ statistic is paradoxically different from a crosstabulation table with more evenly distributed agreements and disagreements, even though the percent of agreement and disagreement do not change. Therefore, the prevalence adjusted bias adjusted kappa (PABAK) will also be reported. PABAK gives the true proportion of agreement beyond expected chance agreement regardless of unbalanced data patterns (28). Both κ and PABAK values were interpreted using Landis and Koch categorization (29) as follows: almost perfect (>0.80), substantial (0.61–0.80), moderate (0.41–0.60), fair (0.21–0.40), slight (0.00–0.20) and poor (<0.00).

We further evaluated both individual and facility level factors that are associated with high level an agreement. To do this we adopted the methods by Morón-Duarte et al (30) to develop a composite score of ANC indicators with adjusted PABAK coefficients in the categories of almost perfect to moderate. In this study, four out of the six indicators were in the categories of almost perfect to moderate as shown in Table 4. We assigned a value of 1 to each indicator agreement (1 = yes/yes or no/no) and 0 otherwise. We multiplied the score by 25 to get a range of values between 0-100. The score was dichotomized into low (≤70 points as 0) and high agreement (≥ 75 points as 1) which was then used to conduct bivariate analysis using the score as the dependent variable. Heterogeneity chi-squared tests were used to measure the difference between low and high agreement categories. All analyses were conducted in Stata 17.

## Results

### Sample characteristics

Table 1 provides characteristics of the women that were available on the day of assessment and had agreed to be both observed and interviewed. Most of the women were 25 years or older (45%). About 14% of the women never attended school, with the majority (61%) reporting to have at least primary education. About 24% of the women were at the antenatal clinic for their first pregnancy. Of the women who came to the facility, 42% was their first time visiting that particular facility. The majority of the women (57%) visited health centers for their ANC visit, and of the facilities they visited, 74% were public facilities. The majority of the ANC consultations were conducted by midwives (75.41%), seconded by nurses (20.60%). Only about 8% of the providers used visual aids in the ANC consultations. Household characteristics such as household wealth were unavailable and therefore not reported.

**Table 1:** Selected social and demographic characteristics of the analysis sample.

| **Variables** | **Percent** | **CI %** |
| --- | --- | --- |
| **Age at last birthday** |  |  |
| 13-19 | 20.26 | (18.41 22.24) |
| 20-24 | 32.95 | (30.67 35.32) |
| 25+ | 46.78 | (44.29 49.31) |
| **Mother's education level** |  |  |
| Never attended school | 13.95 | (12.33 15.75) |
| Primary | 61.31 | (58.86 63.70) |
| Secondary | 21.92 | (19.88 24.10) |
| Tertiary | 2.82 | (2.08 3.80) |
| **Parity** |  |  |
| First pregnancy | 24.40 | (22.38 26.54) |
| **Number of visits for the pregnancy to the facility** |  |  |
| First visit | 42.22 | (39.81 44.66) |
| Second visit | 24.01 | (21.96 26.18) |
| 3+ visits | 33.78 | (31.51 36.12) |
| **Type of facility** |  |  |
| Hospital | 40.01 | (37.47 42.69) |
| Health Center | 56.93 | (54.35 59.48) |
| Dispensary/Clinic/Health post | 3.01 | (2.40 3.78) |
| **Facility managing authority** |  |  |
| Public | 73.65 | (71.59 75.62) |
| Private | 26.35 | (24.38 28.41) |
| **Type of provider** |  |  |
| Doctor | 0.50 | (0.27 0.92) |
| Clinician | 3.49 | (2.84 4.29) |
| Nurse | 20.60 | (18.44 22.93) |
| Nurse Midwife | 75.41 | (73.05 77.63) |
| **Provider used visual aids** |  |  |
| Yes | 7.95 | (6.83 9.22) |

Source: Own derived from the 2013/2014 Service Provision Assessment surveys.

### Characteristics of health facilities where direct observations were conducted

In Table 2 we present the characteristics of the facilities where ANC direct observations and client exit interviews were conducted. About 62% of the facilities were health centers and rural based (76%). Almost 69% of the facilities were government owned. Most of the facilities were located in the southern region (48%) and Central region (42%).

**Table 2:**
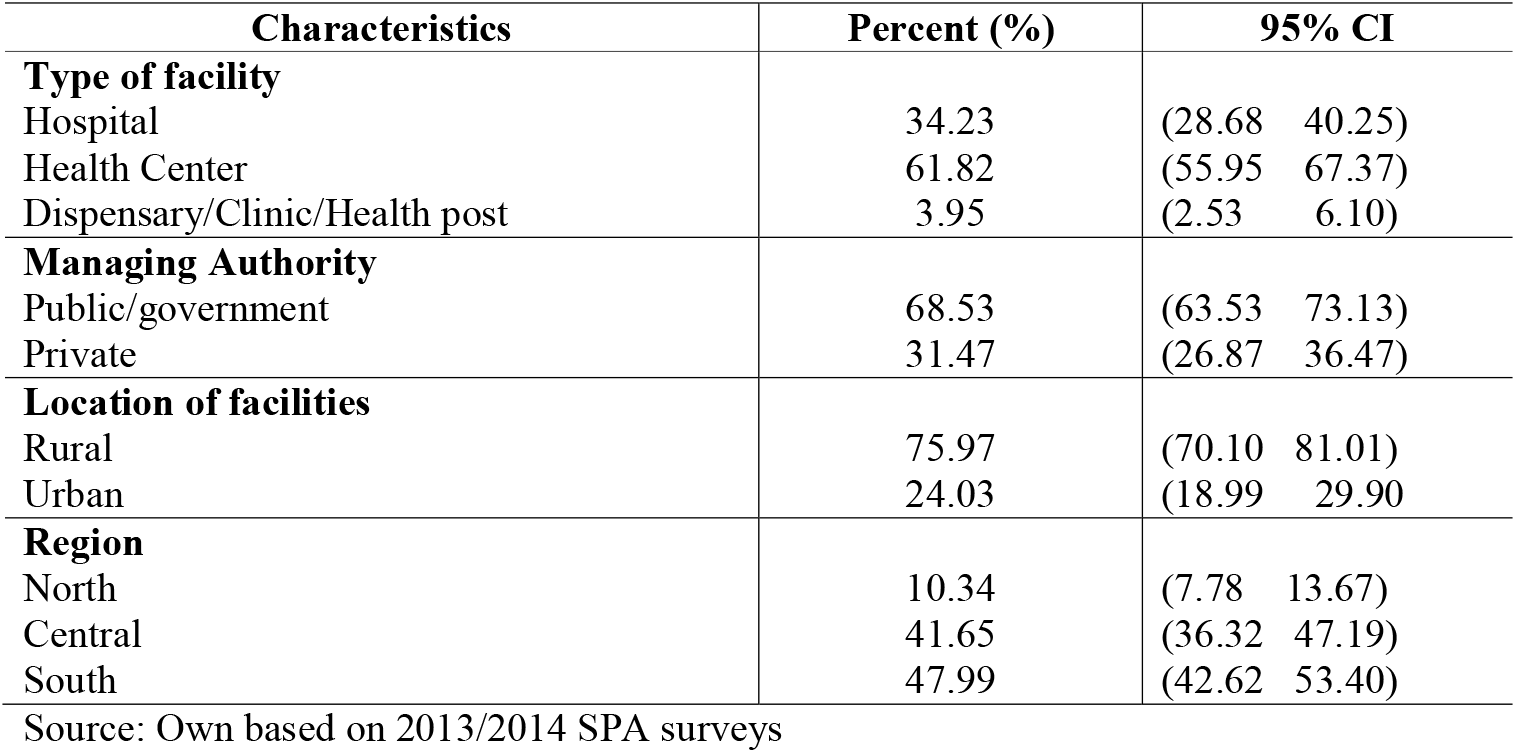
Characteristics of facilities.

### Validation measures

Values for the selected indicators were only missing for less that 1% of the women who respondent “don’t know” (Table 3). The validation estimates show significant variations in specificity and sensitivity across the indicators. The lowest Sensitivity (0.33, 95% CI 0.31-0.35) was observed for whether the woman was informed about the side effects of iron tablets. Whereas the lowest specificity was observed for whether the woman was informed about delivery preparation (0.44, 95%CI 0.42-0.46). In general, the sensitivity of the self-reported exit interviews was high (>90%) for 16.67% of the indicators evaluated, good (≥80 and < 90%) for 33.33% and low (<80%) for 50% of the indicators (all of them in the components of counselling). Specificity was low (<80%) for 66.67% of indicators and high for whether the provider prescribed malaria prophylaxis (84%, 95% CI: 0.82-0.86) and whether the woman was informed of iron tablets side effects (89%, 95% CI: 0.87-0.90).

**Table 3:** Sensitivity and specificity of reporting in exit interviews compared to direct observation in Service Provision Assessment Survey.

| Variable | Client reported (n) | Don't know (n) | N | Client reported prevalence | Observed prevalence | Estimated population prevalence based on sensitivity & specificity | Sensitivity (95% CI) | Specificity (95% CI) | AUC | IF | Met both criteria |
| --- | --- | --- | --- | --- | --- | --- | --- | --- | --- | --- | --- |
| Given or prescribed malaria prophylaxis | 2,068 | 0 | 2,068 | 59.61% | 62% | 59% | 0.86 (0.85 0.88) | 0.84 (0.82 0.86) | 0.84 (0.83 0.86) | 0.96 | Y |
| Given or prescribed iron tablets | 2,068 | 0 | 2,068 | 86.49% | 86.50% | 87.29% | 0.96 (0.95 0.97) | 0.71 (0.69 0.73) | 0.84 (0.81 0.87) | 1 | Y |
| provider ever discussed side effects of iron tablets | 2,050 | 10 | 2,068 | 13.59% | 9.52% | 13.49% | 0.33 (0.31 0.35) | 0.89 (0.87 0.90) | 0.58 (0.55 0.60) | 1.42 | N |
| provider ever discussed diet/nutrition during pregnancy | 2,061 | 7 | 2,068 | 49.89% | 39.14% | 48% | 0.74 (0.72 0.76) | 0.66 (0.64 0.69) | 0.69 (0.67 0.71) | 1.26 | N |
| provider ever discussed preparation for delivery | 2,061 | 7 | 2,068 | 74.86% | 74.63% | 74.63% | 0.80 (0.79 0.82) | 0.44 (0.42 0.46) | 0.62 (0.60 0.65) | 0.99 | N |
| provider talked about danger signs in pregnancy | 2,062 | 6 | 2,068 | 51.36% | 46.30% | 51.51% | 0.61 (0.59 0.63) | 0.56 (0.54 0.59) | 0.59 (0.56 0.61) | 1.11 | N |

**Table 4:** Concordance measures of ANC indicators from the direct observations and client exit interviews.

| Indicator | Concordance (%) |  | Discordance (%) |  | % Agreement<br>(95% CI) | Kappa<br>(95% CI) | P-Value | Agreement<br>strength | PABAK<br>(95% CI) | Agreement<br>strength | P-Value |
| --- | --- | --- | --- | --- | --- | --- | --- | --- | --- | --- | --- |
|  | Y/Y | N/N | Y/N | N/Y |  |  |  |  |  |  |  |
| Given or prescribed malaria prophylaxis | 86.81 | 84.90 | 13.19 | 15.10 | 86.06<br>(84.59 87.58) | 0.71<br>(0.68 0.74) | 0.0000 | Substantial | 0.72<br>(0.69 0.75) | Substantial | 0.0000 |
| Given or prescribed iron tablets | 95.84 | 73.39 | 26.61 | 4.16 | 92.81<br>(91.69 93.92) | 0.69<br>(0.65 0.74) | 0.0000 | Substantial | 0.86<br>(0.83 0.88) | Almost perfect | 0.0000 |
| provider ever discussed side effects of iron tablets | 32.07 | 88.36 | 11.64 | 67.93 | 82.98<br>(81.33 84.64) | 0.17<br>(0.11 0.23) | 0.0000 | Slight | 0.66<br>(0.63 0.69) | Substantial | 0.0000 |
| provider ever discussed diet/nutrition during pregnancy | 74.85 | 66.26 | 33.74 | 25.15 | 69.63<br>(67.64 71.63) | 0.39<br>(0.35 0.43) | 0.0000 | Fair | 0.39<br>(0.35 0.43) | Fair | 0.0000 |
| provider ever discussed preparation for delivery | 81.28 | 44.11 | 55.89 | 18.72 | 71.88<br>(69.92 73.83) | 0.26<br>(0.21 0.30) | 0.0000 | Fair | 0.44<br>(0.40 0.48) | Moderate | 0.0000 |
| provider talked about danger signs in pregnancy | 60.43 | 56.45 | 43.55 | 39.57 | 58.29<br>(56.16 60.43) | 0.17<br>(0.13 0.21) | 0.0000 | Slight | 0.17<br>(0.12 0.21) | Slight | 0.0000 |

Four indicators had AUC results of 0.60 or greater (Table 3 and fig 1). The most accurately reported responses were to the question on whether the provider gave or prescribed malaria prophylaxis (0.84, 95% CI: 0.83–0.86); whether the provider gave or prescribed iron tablets (0.84 95% CI: 0.81-0.87); provider discussed about the importance of good nutrition in pregnancy (0.69, 95%CI: 0.67-0.71) and whether the woman was informed of delivery preparation (0.62, 95% CI: 0.60-065). Indicators with high ROC results mostly reflected objective measures of ANC, where physical care was given (e.g. provision of iron and malaria prophylaxis). Indicators with low value ROC results were mostly subjective measures and required a certain level of knowledge and understanding about the service (counselling on pregnancy complications and iron side effects).

The other criterion of acceptable validity was an inflation factor between 0.75 and 1.25, four indicators met this criterion. These were: the woman was given or prescribed malaria tablets (0.96); was given or prescribed iron tablets (1); the woman was informed of what she needed to do to prepare for delivery (0.99); and the woman was informed of pregnancy related complications (1.11). Two of the indicators with an inflation factor of greater than 1.25 had lower observed prevalence rates in comparison to the other indicators. For example, only 15% of the women, were observed being informed about iron tablets side effects (IF=1.42) and 39% of women were observed being informed about proper nutrition or important foods during pregnancy (IF=1.26). The low observed prevalence explains why the two indicators poor reporting given that even small deviations from 100% in specificity can lead to extreme over-estimation in a survey (Stanton et al. 2013).

### Measures of agreement

Using Kappa estimates, of the six indicators we observed that agreement strength with expert direct observation was slight for two indicators (K between 0.00–0.20), fair for two (K between 0.21–0.40), and substantial for two (K>=0.60) (Table 4). The agreement strength category obtained with the PABAK was higher than that obtained with the Kappa coefficient in three (50%) indicators; whether the woman was prescribed or given iron tablets, whether the woman was informed of the side effects of iron tablets, and whether the woman was informed of what to prepare for delivery. Whereas the other three indicators were equally categorised by PABAK and Kappa values.

### Factors associated with agreement between direct observations and women’s self-reports

In Table 5 we present factors associated with the agreement between the ANC direct observation reports and self-reports in the exit interviews. There was a significantly higher agreement among pregnant women who were younger than 25 years, with primiparous status, those whose ANC visit was the first, those whose health care provider was not a doctor and those whose facilities were located in the central/southern region.

**Table 5:** Factors associated with agreement between ANC direct observation reports and self-reports in the client exit interviews.

| Characteristics | Low agreement (%) | High agreement (%) | P-Value |
| --- | --- | --- | --- |
| **Age category** |  |  |  |
| 13-19 | 10.48 | 89.52 | 0.010** |
| 20-24 | 12.73 | 87.27 |  |
| 25+ | 16.31 | 83.69 |  |
| **Maternal education** |  |  |  |
| Never attended school | 17.02 | 82.98 | 0.257 |
| Primary | 13.27 | 86.73 |  |
| Secondary or higher | 13.98 | 86.02 |  |
| **Parity** |  |  |  |
| First pregnancy | 11.35 | 88.65 | 0.053* |
| ≥ 2 children | 14.80 | 85.20 |  |
| **Number of visits for the pregnancy to the facility** |  |  |  |
| First visit | 8.44 | 91.56 | 0.000*** |
| Second visit | 17.89 | 82.11 |  |
| 3+ visits | 17.91 | 82.09 |  |
| **Region** |  |  |  |
| North | 17.72 | 82.28 | 0.010** |
| Central | 15.14 | 84.86 |  |
| South | 11.49 | 88.51 |  |
| **Type of facility** |  |  |  |
| Hospital | 14.53 | 85.47 | 0.879 |
| Health Center | 13.69 | 86.31 |  |
| Dispensary/Clinic/Health post | 14.58 | 85.42 |  |
| **Facility managing authority** |  |  |  |
| Public | 14.04 | 85.96 | 0.848 |
| Private | 13.72 | 86.28 |  |
| **Type of provider** |  |  |  |
| Doctor | 50.00 | 50.00 | 0.008*** |
| Clinician | 17.02 | 82.98 |  |
| Nurse | 14.01 | 85.99 |  |
| Nurse Midwife | 13.54 | 86.46 |  |

## Discussion

Providing high quality maternal health care services is essential for improving the health and survival of women and new-borns. Accurate information on the care received is therefore, essential to monitoring progress. This study extends prior validation research related to maternal and new-born care interventions by assessing the validity of ANC service coverage indicators using data from the 2013-2014 Malawi service provision assessment (MSPA) which provides information on both direct observations by an expert and women’s report about the service received in the exit interview.

### Validity measures

Our findings show that women’s self-reports in the exit interviews presented overall higher reporting accuracy for indicators related to concrete, observable interventions. For example, indicators which met accuracy criteria reflected those to do with medical prescription (malaria tablets and iron tablets). In contrast, indicators that reflected more abstract concepts, particularly those pertaining to counselling or advice given, performed less reliably. Neither recalling whether the woman was counselled on side effects of iron tablets, diet/nutrition during pregnancy, preparing for delivery and danger signs in pregnancy met both validation criteria. All of these had AUC below the 0.70 benchmark.

Worth to be noted is that despite the lower reporting accuracy, two of the counselling indicators had higher sensitivity than the others; provider ever discussed diet/nutrition during pregnancy (0.74 95% CI: 0.72 -0.76) and provider ever discussed preparation for delivery (0.80, 95%CI: 0.79 0.82). There is a possibility that in these cases counselling was paired with an observable action such as a visual aid showing groups of foods and plastic black paper which is usually recommended as part of delivery in public facilities.

Existing evidence shows mixed findings to the ones reported in this study. In support of this finding is the study by McCarthy et al. (29) in Bangladesh, Cambodia and Kenya. The study has the similar design to this study and found lower reporting accuracy of indicators related to counselling. Similarly, a study by Onishi and Peters (30) conducting a Comparative analysis of exit interviews and direct clinical observations in Paediatric Ambulatory Care Services in Afghanistan found low prevalence of counselling items (ranging from 8 to 80%). The authors also found that exit interviews had relatively low levels of sensitivity for the counselling items, ranging from 33 to 88%, with higher levels of specificity (ranging from 63 to 91%), whereas the ROCs ranged from 61 to 77% (30).

In Brazil, Morón-Duarte et al (31) examined the agreement of antenatal care indicators from self-reported questionnaire and the antenatal care card also reported low reporting accuracy of counselling indicators. On the other hand, a recall study of maternal and newborn interventions received in the postnatal period in Kenya and eSwatini found that some indicators of physical examination had higher reporting accuracy (32). However, the study also found somewhat better recall of counselling indicators including whether the provider discussed danger signs for the mother. Another study in China which compared women’s reports with facility records found generally lower specificity for indicators of the maternal physical examination during ANC and PNC and only prior known PNC validation study (33).

### Reliability or measures of agreement

We see the similar trend that observable interventions such as drug prescription had moderate to high agreement. Suggesting that assessment of ANC through self-reports is reliable and the same as using direct observation for those specific components. These findings may be related to the ability of the patient to identify the reason for the ANC procedure/action recorded by the health professional during the ANC visit. Whereas indicators on counselling had moderate to slight agreement except for one which had substantial agreement (a woman’s recall on whether she got counselled on side effects of iron tablets). In general, counselling indicators had lower prevalence rates in comparison to the observable interventions (see table 3), therefore, the low agreement may be because the counselling is not provided as it is recommended.

These findings are similar to those found by Assaf et al. (20) that ANC counselling indicators had low agreement in Haiti, Malawi and Senegal. Worth to be noted however is that the Assaf study only focused on the counselling aspect of maternal and child care interventions and did not include the observable components of ANC. In Brazil, Morón-Duarte et al. (31) also reported moderate to high agreement on indicators of service utilization, clinical examination and diseases during pregnancy but poor agreement on indicators of counselling. Bessinger and Bertrand (18) conducted a study in Ecuador, Uganda and Zimbabwe to examined the comparability of reports from direct observations and exit interviews at health facilities. The authors found that for the majority of indicators, agreement was good to excellent and was relatively higher on the indicators of interpersonal relationships but lower on those related to counselling.

### Factors associated with agreement between client interviews and ANC direct observations

Individual sociodemographic characteristics as well as facility characteristics may contribute towards the strength of agreement between data sources (31, 34). In this study, we found that at an individual level, women who were younger than 25 years, with primiparous status and those whose ANC visit was the first had significantly higher agreement. A possible explanation could be because of their perceived low experience, women in these categories are likely to be more attentive and seek clarification where possible unlike women who are older or have other kids may think they already know so much from experience and miss out on the new information from the provider (31). A study by Morón-Duarte et al.(31) reports similar findings that higher maternal age and pregnant women with ≥2 children were associate with a lower probability of high agreement between the antenatal card and the self-reported questionnaire.

We also found that women whose consultations were done by doctor had low agreement compared to those whose ANC consultations were conducted by clinicians, nurses or midwives. This is not a surprising finding as a Cochrane systematic review found that care delivered by nurses, compared to care delivered by doctors, probably generates similar or better health outcomes for a broad range of patient conditions (35). Lastly, we did also observe high agreement in facilities located in the central/southern region. For the most part, we believe that the issue could be language. In the northern region of Malawi, Tumbuka is the official language and Chichewa is not as common as it is in the Southern and Central regions. There is a possibility that much of the low agreement in the Northern region was due to the language barrier between the provider and the client especially in cases where the provider was not a native speaker of Tumbuka. The issue of language has to be explored more to assess its impact on understanding and information retention during clinical consultation.

### Limitations and study implications

The major strength of this study is the use of facility level census data and the use of direct observations as a gold standard. On the other hand, the interpretation of these results should be done with caution. First, our analysis was limited to only indicators which were available in both direct observation guide and client exit interview questionnaire which do not represent overall process of ANC. Second, while direct observation by an expert is considered to be the gold standard, it may also be imperfect. Differences in observer training protocols, facility practices or how apparent it was that a given intervention was implemented (especially the counselling components), among other factors, may contribute to differences in observer ratings across settings (19). Finally, an important consideration of the relevance of these study findings for national and global monitoring efforts is that this study assessed women’s immediate recall accuracy (at facility discharge). Results may therefore, not be directly generalisable to the DHS and MICS, which have longer recall periods.

## Conclusion

Our study shows that data from women’s self-reports provide substantially different information on indicator performance from direct observation especially on counselling related indicators. From a public health perspective this raises questions with regard to reliance on household surveys such as MICS and DHS which have a longer recall period in monitoring quality of healthcare services. While immediate recall may represent best-case scenario in terms of accuracy, findings inform the degree to which women perceived specific interventions took place. Prior evidence suggests that, unless interventions are recalled with high accuracy at facility discharge, recall generally declines with time. For example, validation analysis of facility-based interventions received in the intrapartum and immediate postnatal periods in Kenya showed that the few select interventions which were recalled with high accuracy at facility discharge maintained acceptable accuracy at 13–15 months’ follow-up (8). However, for most indicators, recall accuracy was poor and either remained the same or declined with time.

While additional research evaluating different lengths of recall time is warranted, it is possible that immediate recall is necessary in order to ‘code’ certain events into memory for later reporting. In the context of calls for enhanced measurement of the components that lead to effective coverage, study findings such as these suggest that careful consideration of the type of information women are asked to recall is needed. While household survey programmes such as the DHS and MICS are frequently relied on as data sources for measuring intervention coverage, triangulation of such findings with other data sources such as client exit interviews is important.

## Data Availability

https://dhsprogram.com/methodology/survey/survey-display-424.cfm

## Footnotes

1 Adapted from (Munos et al. 2018).

## Notes

### Competing Interest Statement

The authors have declared no competing interest.

### Funding Statement

The authors received no specific funding for this work

